# Challenges in the implementation of preventive treatment for tuberculosis in people living with HIV/Aids: A qualitative study

**DOI:** 10.1101/2023.09.12.23295456

**Authors:** Vânia Silva dos Reis, Débora Dupas Gonçalves do Nascimento, Terezinha Alcântara da Silva, Anamaria Mello Miranda Paniago, Adriana Carla Garcia Negri, Gabriela Ferreira, Rafaela Ferreira, Everton Ferreira Lemos, Anete Trajman, Sandra Maria do Valle Leone de Oliveira

## Abstract

Treatment of tuberculosis infection (TBI) in people living with HIV/Aids (PLWHA) reduces the risk of tuberculosis disease (TBD), the main cause of death in this population. Recognizing the barriers related to the scale up of tuberculosis preventive treatment (TPT) can contribute to reorganization of health services for achieving TB elimination. This qualitative study aimed to understand the perception of specialized care health professionals of a capital city in Brazil about TPT for PLWHA. Between October 2020 and August 2022 ten physicians and four nurses working for more than six months in reference HIV/Aids services were interviewed. Interviews were guided by a script of previously validated, semi-structured questions. The audios were recorded, transcribed, and categorized based on Grounded Theory, and its analysis was anchored in the theoretical framework of Symbolic Interactionism. Sampling was performed using theoretical saturation. The emerged central phenomenon “Facing challenges in the implementation of TPT in the light of scientific evidence” was supported by four categories:1) Demonstrating theoretical knowledge regarding TBI and its treatment guidelines; 2) Dealing with the complexities of guidelines adherence in the routine service; 3) Building bonds to overcome different challenges in the care of PLWHA; 4) Seeking strategies to facilitate adherence to the TPT guidelines service. Professionals faced challenges in implementing TPT due to structural obstacles in service, hesitation in establishing therapy, and the risk of adverse events when dealing with patients’ vulnerabilities, despite available scientific evidence, theoretical knowledge, and guidelines recommendations. They envisaged ideal conditions to adhere to the TPT guidelines in service, and seeked to mediate confrontations by strengthening their bond with patients through multidisciplinary interaction, teaching, and research.

## Introduction

Tuberculosis preventive treatment (TPT) is a crucial component of the strategy devised to end the global tuberculosis (TB) epidemic [1–4]. Although preventable and curable, TB is still the leading cause of death among people living with HIV/Aids (PLWHA), accounting for one-third of the deaths [1].

TPT in PLWHA was one of the top priorities at the high-level meeting of the United Nations (UN) on TB in 2018 [4] and globally, the target of offering 6 million TPT for PLWHA was attained and surpassed in 2021 [5]. This was achieved based on the World Health Organization (WHO) recommendation to offer TPT to PLWHA regardless of TBI tests and chest radiography (CXR) to exclude disease tuberculosis (TBD) when these tests were unavailable [1].

However, TPT uptake in PLWHA and other high-risk populations is low in Brazil, one of the 30 countries with high burden of TB and TB/HIV co-infection [1]. Diagnosis and treatment of tuberculosis infection (TBI) are offered free of charge by the Public Unified Health System (SUS) in the country [6], but several challenges lie in the implementation of TPT guidelines, with resulting low rates of TPT initiation and completion [3,7].

Previous studies have shown that educational activities for health care providers in the country can increase TPT prescription among household contacts in SUS, despite the need for testing [7]. This intervention was affordable [8] and sustainable [9]. However, among PLWHA, recent studies have shown low adherence by health professionals in prescribing TPT [10,11].

Understanding the subjectivity of individuals involved in TPT scale up can contribute to adherence to the national guidelines, with adoption of strategies to reduce barriers in health services. The objective of the current study was to understand the perception of specialized care health professionals regarding TPT for PLWHA in a capital city of the Midwest region of Brazil.

## Study design

### Type of study

In this qualitative study [12,13], we used the Grounded Theory (GT) as a methodological resource to guide data collection, analysis, and identification of the phenomenon [13]. Results were analyzed based on Brazilian TBI treatment guidelines, which recommended, at the time of data collection, TST or IGRA for PLWHA with CD4 count >350 cells/mm^3^, those with lower levels TPT is recommended without TBI testing, CXR for all before prescription, and isoniazid as the preferred regimen [6,14]. It was anchored in the theoretical conceptions of Symbolic Interactionism (SI), where perspectives are socially created and changed according to groups and/or roles. SI facilitates the understanding that the process of interaction with others enables an individual to acquire new perspectives, and these guide their way of acting [15]. We used the Standards for Reporting Qualitative Research (SRQR) guidelines to report this study [16,17].

### Participant selection

Inclusion criteria was being a medical or nurse professional working for more than six months in HIV/Aids reference services and agreeing to participate by signing the informed consent. There were no exclusion criteria.

### Data collection

Data collection was carried out in two public institutions of reference in HIV/Aids in the capital of Mato Grosso do Sul, located in the Midwest region of Brazil, between October 2020 and August 2022.

For data collection, a semi-structured interview [18] was used, guided by questions related to the knowledge and perception of professionals about TBI and TPT guidelines for PLWHA, and the bond between patient and service professionals. This script was originally validated by a medical professional and a nurse and, as there was no need for changes, they were included in the study sample. The interviews were recorded in audio by a psychologist and lasted an average of 15 minutes each. The interruption in data collection occurred due to the saturation of theoretical data when the emerged data became repetitive [19,20].

### Data analysis

The collected data were transcribed in full and analyzed according to GT through coding in three stages: open, axial, and selective. In open coding, data were separated, compared, and analyzed line-by-line in detail, generating 568 conceptual codes, which were later grouped by similarities and differences. In axial coding, the data were regrouped, clarifying explanations regarding the phenomena under investigation and allowing the emergence of categories. This regrouping followed the conditional/consequential paradigm composed of three components: condition, action-interaction and consequence. In selective coding, comparison and constant analysis were performed, refining the relationships between the four categories and ten subcategories, integrating them, and seeking to identify one central phenomenon [13,19].

## Results

Fourteen participants were included, 10 were physicians and four were nurses. Of these, 10 were women, nine were married, and 14 lived in urban areas. Their mean age was 45 years. They had an average of 13 years of experience in HIV/Aids reference services, and an average of 22 years of training. Seven of the study participants had a doctorate, nine had a master degree, and 13 were specialized professionals.

The health professional (HCW) demonstrated a reasonable understanding of the concept of TBI and the TPT guidelines for PLWHA. They viewed the guidelines as an essential strategy for reducing deaths and transmission within the community. This perception is evident in the statements made by the participants in Table 1.

**Table 1:**
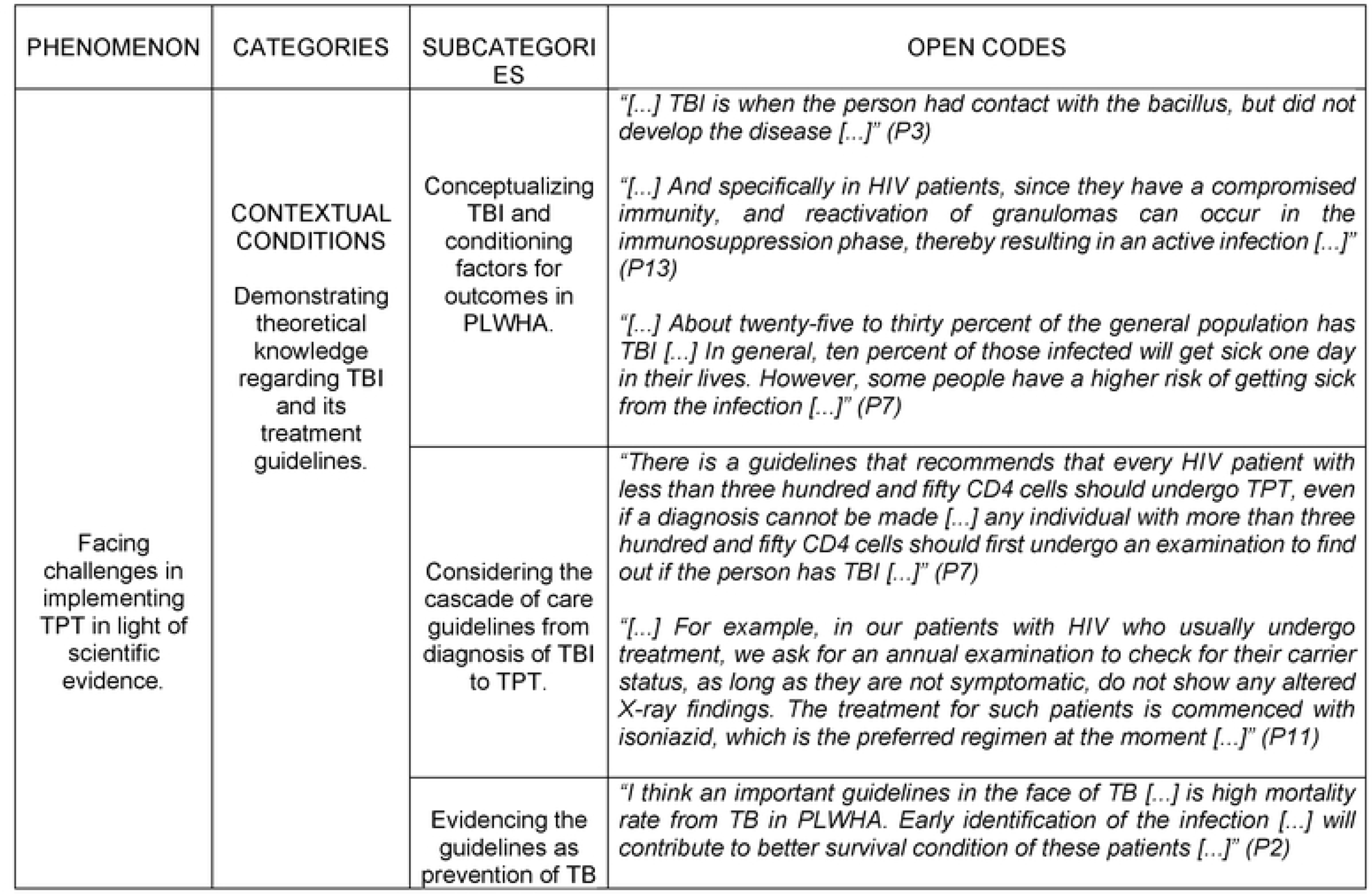
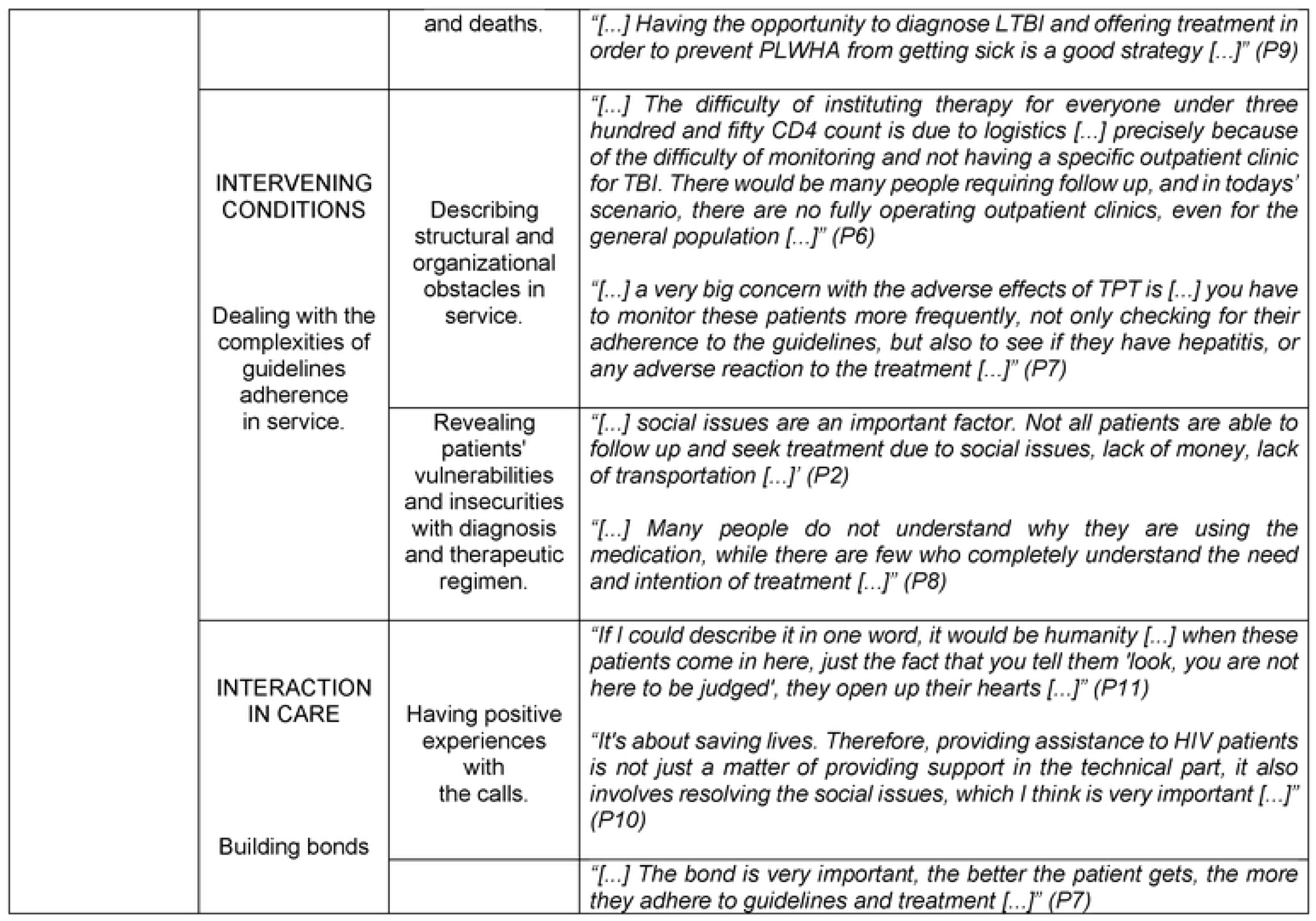
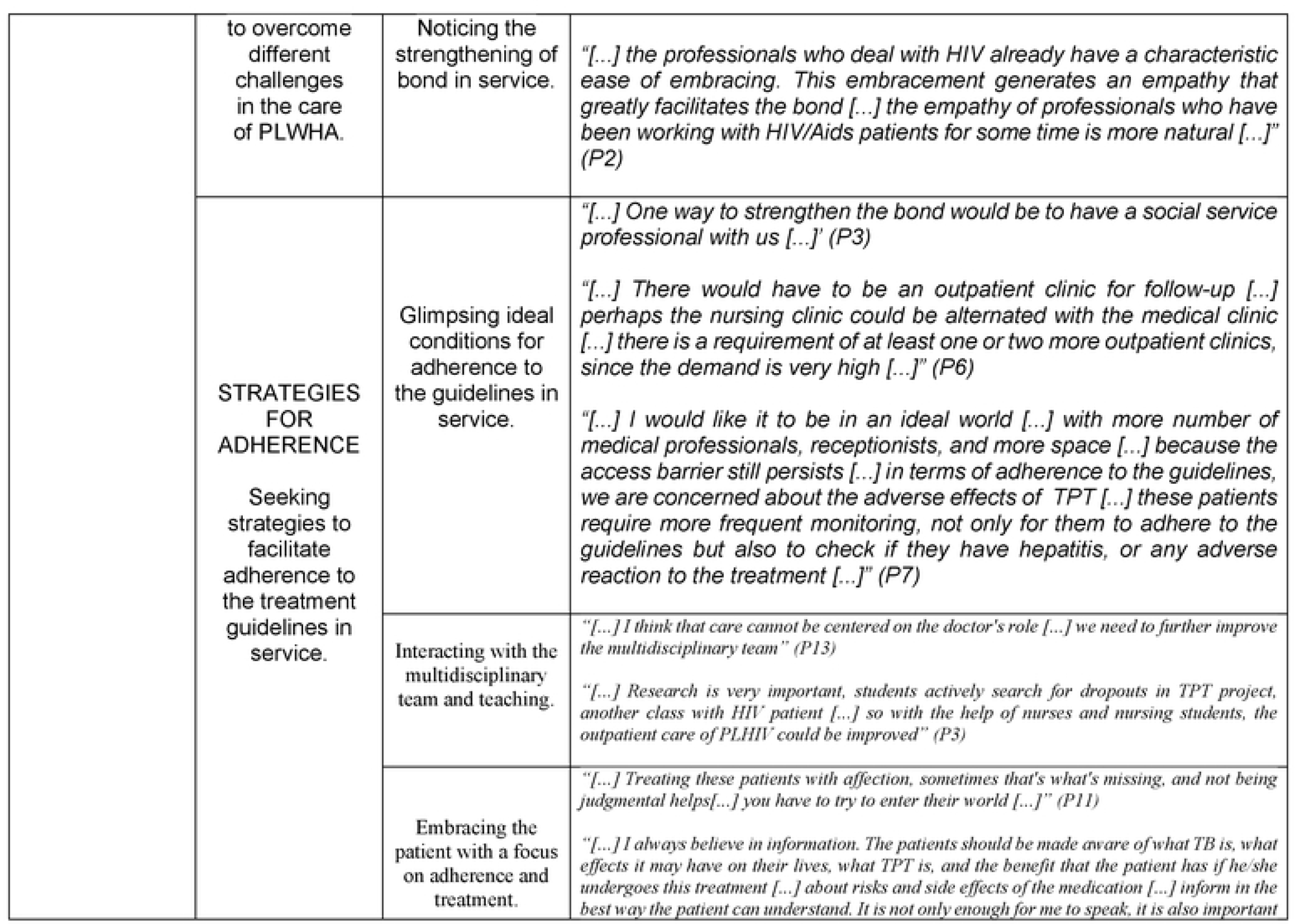
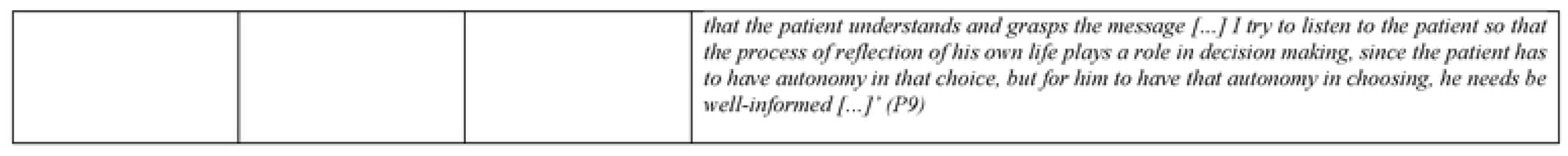
Phenomenon, categories, subcategories and open codes that emerged from the interviews.

However, the HCW also acknowledged the challenges associated with adhering to in-service guidelines. They specifically expressed concerns regarding patient vulnerability, which further complicates the issue of TPT adherence for medical professionals.

In this context, they build bonds to overcome different challenges in care for PLWHA (Table 1):

*“It’s about saving lives. Therefore, providing assistance to HIV patients is not just a matter of providing support in the technical part, it also involves resolving the social issues, which I think is very important […]” (P10)*

*“[…] The bond is very important, the better the patient gets, the more they adhere to guidelines and treatment […]” (P7)*

And seeking strategies to facilitate adherence to the treatment guidelines in service:

*“[…] There would have to be an outpatient clinic for follow-up […] perhaps the nursing clinic could be alternated with the medical clinic […] there is a requirement of at least one or two more outpatient clinics, since the demand is very high […]” (P6)*

Interacting with the multidisciplinary team and teaching:

*“[…] I think that care cannot be centered on the doctor’s role […] we need to further improve the multidisciplinary team” (P13)*

Embracing the patient with a focus on adherence and treatment:

*“[…] I always believe in information. The patients should be made aware of what TB is, what effects it may have on their lives, what TPT is, and the benefit that the patient has if he/she undergoes this treatment […] about risks and side effects of the medication […] inform in the best way the patient can understand. It is not only enough for me to speak, it is also important that the patient understands and grasps the message […] I try to listen to the patient so that the process of reflection of his own life plays a role in decision making, since the patient has to have autonomy in that choice, but for him to have that autonomy in choosing, he needs be well-informed […]’ (P9)*

This study sheds light on the perception of HCW regarding their knowledge and meaning of TPT in PLWHA, challenges faced due to weaknesses of the services, and the strategies that can be used to overcome these limitations.

## Discussion

Knowledge of HCW contrasted with hesitations and uncertainties due to structural limitations of health facilities, but empathy, embracement and multidisciplinary interaction were perceived as enablers for quality care of PLWHA. Professionals perceive and value the bond strengthened in the service and use it as an instrument to overcome structural limitations in the care of PLWHA. Feelings, such as satisfaction, humanization, and empathy, were found to be facilitators of patient’s attachment to the service. These findings were in accordance with SI conceptions, where people modulate their behavior based on the meaning they attribute to things and the social interaction established with others. People modify this meaning through interpretive process they use when faced with things and situations throughout their lives [15].

Knowledge of HCW regarding TBI and its management, along with the strategic importance of preventing TB and unfavorable outcomes in PLWHA have been previously reported [21–24].

Weaknesses in structural and organizational conditions of the services emerged as factors hindering the scale up of TBI treatment. This included other care demands in face of understaffed teams. Lack of training, irregularities in screening and during TPT, ill-defined responsibilities in prescription and record keeping, clinical misperception [25], lack of medication, fear of creating bacterial resistance, low acceptance of medication by patients, side effects, and lack of commitment from health managers have been identified as barriers to implementation of TPT [2]. A recent study in Brazil showed that educational interventions for HCW about TPT and its benefits were fundamental for the increase in treatment prescriptions [9]. Continuous efforts are necessary to improve TPT care among PLWHA [3].

Concerns regarding adverse reactions have emerged as a hindrance in the to TPT and in HCW team adherence. This can be minimized with regular monthly clinical follow-up and clear guidelines for patients [26]. Expansion of human resources and operational research [7], along with adequate economic perspectives for monitoring adverse events [27], have been indicated as strategies to overcome this issue. Still, the fear of prescribing TPT can be resolved with the use of short regimens which have shown greater safety and rare adverse events [28].

Patient vulnerability is also a concern for medical professionals regarding adherence to TPT. The relationship between vulnerabilities and Aids is bidirectional and is strongly influenced by poverty and other markers of social exclusion. Different aspects linked to PLWHA reinforce and produce new vulnerabilities arising from the stigma of the disease, which impacts people’s health even after four decades since the discovery of the disease [29]. Patients’ specific needs must be addressed considering the local context, specific settings and conditions in which the TPT program would be implemented [30].

Combining communication skills and empathy is important for the establishment of satisfactory interpersonal relationships [31]. Strengthening this bond is necessary to empower the patient to face the stigma of the disease, and to favor adherence to treatment [6,32,33]. Facilitation of access and bonding, promptness of care, health professional and patient relationship, duration of appointments, language used, and availability of references for necessary referrals are fundamental requirements of services to promote adherence to treatment [6]. Intensification of strategies for adherence to HIV/TB co-infection prevention treatment is a priority [34].

This study has a few limitations. It was conducted in a single city, in a country with over 5000 municipalities’, and the perspective of HCW may be different according to the multiple existing scenarios. In addition, it is restricted to the HCW perspective. The users’ perspective is equally important to understand the dropouts in the TPT cascade of care. However, there is a paucity of qualitative studies on TPT in PLWHA, and this study clearly identified the central phenomenon “Facing challenges in the implementation of TPT in the light of scientific evidence”, and the strategies to overcome barriers.

## Ethics statement

This study was conducted in accordance with the tenets of Declaration of Helsinki. It was pproved by the Ethics Committee for Research with Human Beings of the Federal University of Mato Grosso do Sul on November 2, 2020 (# 4.375.024). Written informed consent was obtained from all the study participants. To maintain secrecy and anonymity, the participants were represented by the letter “P” followed by the number corresponding to the order of the interviews (P1, P2, P3 …).

## Author Contributions

VSR, DPGN and SMVLO were responsible for the design and conduct of the study, data analysis, writing and review of the article. VSR contributed with the transcription of the interviews. TAS carried out the interviews, initial analysis and review of the article. GF, RF contributed with the transcription of interviews, initial categorization of data, writing and formatting of the article. ACGN, AMMP, EFL assisted in data analysis and article review. AT contributed to the data analysis and review of the article. All authors approved the submitted version.

## Data Availability Statement

The raw data supporting the conclusions of this article will be made available by the authors, without undue reservation.

## Acknowledgments

To all the nurses and physicians, from the reference services in HIV/Aids, who contributed as participants in this study.

## Reference

1. World Health Organization (WHO). Global Tuberculosis Report 2021. Geneva: WHO; 2021. Available from: https://www.who.int/teams/global-tuberculosis-programme/tb-reports/global-tuberculosis-report-2021

2. Teklay G, Teklu T, Legesse B, Tedla K, Klinkenberg E. Barriers in the implementation of isoniazid preventive therapy for people living with HIV in Northern Ethiopia: a mixed quantitative and qualitative study. BMC Pub Health. 2016 Aug; 16(1):840. Available from: https://www.ncbi.nlm.nih.gov/pmc/articles/PMC4992328/

3. Bastos ML, Melnychuk L, Campbell JR, Oxlade O, Menzies D. The latent tuberculosis cascade-of-care among people living with HIV: A systematic review and meta-analysis. PLoS Med. 2021 Sep; 18(9):e1003703. Available from: https://www.ncbi.nlm.nih.gov/pmc/articles/PMC8439450/

4- World Health Organization. Global tuberculosis report 2019. Geneva: WHO; 2019. Available from: https://apps.who.int/iris/bitstream/handle/10665/329368/9789241565714-eng.pdf?ua=1

5- World Health Organization. Global Tuberculosis Report 2022. Geneva: WHO; 2022. Available from: https://www.who.int/teams/global-tuberculosis-programme/tb-reports/global-tuberculosis-report-2022

6- Ministério da Saúde (Brasil), Secretaria de Vigilância em Saúde, Departamento de Vigilância das Doenças Transmissíveis. Manual de Recomendações para o Controle da Tuberculose no Brasil 3a. ed. Brasília: Ministério da Saúde; 2019. 364 p. Available from: https://bvsms.saude.gov.br/bvs/publicacoes/manual_recomendacoes_controle_tuberculose_brasil_2_ed.pdf

7- Bastos ML, Oxlade O, Benedetti A, Fregonese F, Valiquette C, Lira SCC, Carvalho-Cordeiro D, Cavalcante JR, Faerstein E, Albuquerque MFM, Cordeiro-Santos M, Hill PC, Menzies D, Trajman A. A public health approach to increase treatment of latent TB among household contacts in Brazil. Int J Tuberc Lung Dis. 2020 Oct 1;24(10). Available from: https://www.ingentaconnect.com/content/iuatld/ijtld/2020/00000024/00000010/art00005;jsessionid=6oh70t2e9b6cn.x-ic-live-02#

8- Bastos ML, Oxlade O, Campbell JR, Faerstein E, Menzies D, Trajman A. Scaling up investigation and treatment of household contacts of tuberculosis patients in Brazil: a cost-effectiveness and budget impact analysis. Lancet Reg Health Am. 2022 Jan; 10(8). Available from: https://www.ncbi.nlm.nih.gov/pmc/articles/PMC9903685/

9- Yanes-Lane M, Trajman A, Bastos ML, Oxlade O, Valiquette C, Rufino N, Fregonese F, Menzies D. Effects of programmatic interventions to improve the management of latent tuberculosis: a follow up study up to five months after implementation. BMC Public Health. 2021 Jan; 21(1). Available from: https://www.ncbi.nlm.nih.gov/pmc/articles/PMC7819253/

10- Sobral L, Arriaga MB, Souza AB, Araújo-Pereira M, Barreto-Duarte B, Sales C, Rocha MS, Benjamin A, Moreira ASR, de Oliveira JG, Carvalho AC, Spener-Gomes R, Figueiredo MC, Cavalcante S, Durovni B, Lapa-E-Silva JR, Kritski AL, Rolla VC, Sterling TR, Cordeiro-Santos M, Andrade BB. Determinants of losses in the tuberculosis infection cascade of care among children and adolescent contacts of pulmonary tuberculosis cases: A Brazilian multi-centre longitudinal study. Lancet Reg Health Am. 2022 Nov;15:100358. doi: 10.1016/j.lana.2022.100358. Epub 2022 Aug 23. PMID: 36438860; PMCID: PMC9696515. Available from: https://pubmed.ncbi.nlm.nih.gov/36438860/

11- Alsdurf H, Hill PC, Matteelli A, Getahun H, Menzies D. The cascade of care in diagnosis and treatment of latent tuberculosis infection: a systematic review and meta-analysis. Lancet Infect Dis. 2016 Nov;16(11):1269–78. Available from: https://pubmed.ncbi.nlm.nih.gov/27522233/

12- Minayo, M. C. S. Ética das pesquisas qualitativas segundo suas características. Revista Pesquisa Qualitativa. 2021 Dez; 9(22): 521–539. Available from: https://editora.sepq.org.br/rpq/article/view/506/290

13- Corbin J, Strauss A. Basics of qualitative research: techniques and procedures for developing Grounded Theory. California: SAGE; 2015.

14- Ministério da Saúde (Brasil), Secretaria de Vigilância em Saúde, Departamento de Vigilância das Doenças Transmissíveis. Protocolo de vigilância da infecção latente pelo Mycobacterium tuberculosis no Brasil. 1a. ed. Brasília: Ministério da Saúde; 2018. 32 p. Available from: https://bvsms.saude.gov.br/bvs/publicacoes/protocolo_vigilancia_infeccao_latente_mycobacterium_tuberculosis_brasil.pdf

15- Blumer H. Symbolic interacionism: perspective e method. Berkeley (US): University of Califórnia; 1969.

16- O’brien BC, Harris IB, Beckman TJ, Reed DA, Cook DA. Standards for reporting qualitative research: a synthesis of recommendations. Acad Med. 2014 Sep; 89(9):1245–51. Available from: https://pubmed.ncbi.nlm.nih.gov/24979285/

17- Saab MM, O’driscoll M, Fitzgerald S, Sahm LJ, Leahy-Warren P, Noonan B; et al. Primary healthcare professionals’ perspectives on patient help-seeking for lung cancer warning signs and symptoms: a qualitative study. BMC Prim Care. 2022 May; 23(1):119. Available from: https://www.ncbi.nlm.nih.gov/pmc/articles/PMC9114293/pdf/12875_2022_Article_1730.pdf

18- Moser A, Korstjens I. Series: Practical guidance to qualitative research. Part 3: Sampling, data collection and analysis. Eur J Gen Pract. 2018 Dec; 24(1):9–18. Available from: https://www.ncbi.nlm.nih.gov/pmc/articles/PMC5774281/

19- Strauss A, Corbin J. Pesquisa Qualitativa: técnicas e procedimentos para o desenvolvimento de teoria fundamentada. 2a. ed. Porto Alegre: Artmed; 2008.

20- Minayo MCS, Costa AP. Fundamentos Teóricos das Técnicas de Investigação Qualitativa. Revista Lusófona de Educação. 2018; 40(1):139–53. Available from: file:///C:/Users/user/Downloads/6439-Texto%20do%20artigo-19398-1-10-20180827%20(2).pdf

21- Macneil A, Glaziou P, Sismanidis C, Date A, Maloney S, Floyd K. Global Epidemiology of Tuberculosis and Progress Toward Meeting Global Targets - Worldwide, 2018. MMWR Morb Mortal Wkly Rep. 2020 Mar; 69(11):281–85. Available from: https://www.ncbi.nlm.nih.gov/pmc/articles/PMC7739980/

22- Picone CM, Freitas AC, Gutierrez EB, Silva VIA. Acesso e adesão à terapia preventiva com isoniazida e ocorrência de TB ativa em uma coorte de pessoas vivendo com HIV: um estudo de coorte retrospectivo em São Paulo, Brasil. Revista do Instituto de Medicina Tropical de São Paulo. 2020; 62(8). Available from: https://www.scielo.br/j/rimtsp/a/XgVGv9MCnTMKCZ89S5kxrbH/abstract/?lang= en#

23- Hasan T, Au E, Chen S, Tong A, Wong G. Screening and prevention for latent tuberculosis in immunosuppressed patients at risk for tuberculosis: a systematic review of clinical practice guidelines. BMJ Open. 2018 Set; 08(9):01–14. Available from: https://www.ncbi.nlm.nih.gov/pmc/articles/PMC6144320/pdf/bmjopen-2018-022445.pdf

24- Oxlade O, Trajman A, Benedetti A, Adjobimey M, Cook VK, Fisher D.; et al. Enhancing the public health impact of latent tuberculosis infection diagnosis and treatment (ACT4): protocol for a cluster randomised trial. BMJ Open. 2019 Jan. Available from: https://bmjopen.bmj.com/content/bmjopen/9/3/e025831.full.pdf

25- Roscoe C, Lockhart C, De Klerk M, Baughman A, Agolory S, Gawanab M; et al. Evaluation of the uptake of tuberculosis preventative therapy for people living with HIV in Namibia: a multiple methods analysis. BMC Public Health 20. 2020 Apr; 1838. Available from: https://bmcpublichealth.biomedcentral.com/articles/10.1186/s12889-020-09902-z#citeas

26- Getahun H, Matteelli A, Abubakar I, Aziz MA, Baddeley A, Barreira D; et al. Management of latent Mycobacterium tuberculosis infection: WHO guidelines for low tuberculosis burden countries. Eur Respir J. 2015 Dec; 46(6):1563–76. Available from: https://www.ncbi.nlm.nih.gov/pmc/articles/PMC4664608/

27- Sotgiu G, Matteelli A, Getahun H, Girardi E, Sañé Schepisi M, Centis R; et al. Monitoring toxicity in individuals receiving treatment for latent tuberculosis infection: a systematic review versus expert opinion. Eur Respir J. 2015 Apr; 45(4):1170–3. Available from: https://www.ncbi.nlm.nih.gov/pmc/articles/PMC4391659/

28- Sterling TR, Njie G, Zenner D, Cohn DL, Reves R, Ahmed A, Menzies D, Horsburgh CR Jr, Crane CM, Burgos M, LoBue P, Winston CA, Belknap R. Guidelines for the Treatment of Latent Tuberculosis Infection: Recommendations from the National Tuberculosis Controllers Association and CDC, 2020. MMWR Recomm Rep. 2020 Feb 14;69(1):1–11. doi: 10.15585/mmwr.rr6901a1. PMID: 32053584; PMCID: PMC7041302. Available from: https://www.ncbi.nlm.nih.gov/pmc/articles/PMC7041302/

29- Damião JJ, Rafael A, Maksud I, Filgueiras S, Rocha F, Maia AC; et al. Cuidando de Pessoas Vivendo com HIV/Aids na Atenção Primária à Saúde: nova agenda de enfrentamento de vulnerabilidades? Saúde em Debate. 2022; 46(132):163–74. Available from: https://www.scielo.br/j/sdeb/a/XqmLCJ7cLZg94kp8DjjqKSy/#

30- Stuurman AL, Vonk NSM, Van Kessel F, Oordt-Speets AM, Sandgren A, Van Der Werf MJ. Interventions for improving adherence to treatment for latent tuberculosis infection: a systematic review. BMC Infect Dis. 2016 Jun; 8(16):257. Available from: https://www.ncbi.nlm.nih.gov/pmc/articles/PMC4897858/

31- Gularte NDG, Velho MTAC, Gonçalves KCS; Beschoren NF. A Relação Clínica e a Comunicação de Notícias Difíceis com o Auxílio das Artes e dos Relatos Vivos. Abordando a Relação Clínica e Comunicando Notícias Difíceis com o Auxílio das Artes e das Histórias Vivas. Rev Bras Educ Med. 2019; 43(4):131–40. Available from: https://www.ncbi.nlm.nih.gov/pmc/articles/PMC8087288/

32- Da Silva RD, De Luna FDT, De Araújo AJ, Camêlo ELS, Bertolozzi MR, Hino P; et al. Patients’ perception regarding the influence of individual and social vulnerabilities on the adherence to tuberculosis treatment: a qualitative study. BMC Public Health. 2017 Sep; 17(1):725. Available from: https://www.ncbi.nlm.nih.gov/pmc/articles/PMC5606083/

33- Souza LLL, Santos FLD, Crispim JA, Fiorati RC, Dias S, Bruce ATI; et al. Causes of multidrug-resistant tuberculosis from the perspectives of health providers: challenges and strategies for adherence to treatment during the COVID-19 pandemic in Brazil. BMC Health Serv Res. 2021 Oct; 21(1):1033. Available from: https://www.ncbi.nlm.nih.gov/pmc/articles/PMC8483800/

34- Ministério da Saúde (Brasil), Secretaria-Executiva. Recomendações para o manejo da coinfecção TB-HIV em serviços de atenção especializada a pessoas vivendo com HIV/AIDS. 1a. ed. Brasília: Ministério da Saúde; 2013. 28 p. Available from: https://bvsms.saude.gov.br/bvs/publicacoes/recomendacoes_manejo_coinfeccao_tb_hiv.pdf

